# The impact of blinding on trial results: A systematic review and meta-analysis

**DOI:** 10.1101/2023.03.05.23286821

**Authors:** Tyler Pitre, Sarah Kirsh, Tanvir Jassal, Mason Anderson, Adelia Padoan, Alexander Xiang, Jasmine Mah, Dena Zeraatkar

## Abstract

**Background:** Blinding—the concealment of the arm to which participants have been randomized—is an important consideration for assessing risk of bias of randomized trials. A growing body of evidence has, however, yielded inconsistent results on whether trials without blinding produce biased findings.

**Objective:** To conduct a systematic review and meta-analysis of the evidence addressing whether trials with and without blinding produce different results.

**Methods:** We searched MEDLINE, EMBASE, Cochrane Reviews, JBI EBP, and Web of Science, from inception to May 2022, for studies comparing the results of trials with and without blinding. Pairs of reviewers, working independently and in duplicate, reviewed search results for eligible studies and extracted data.

We pooled the results of studies comparing trials with and without blinding of patients, healthcare providers/investigators, and outcome assessors/adjudicators using frequentist random-effects meta-analyses. We coded study results such that a ratio of odds ratio (ROR) < 1 and difference in standardized mean difference (dSMD) < 0 indicate that trials without blinding overestimate treatment effects.

**Results:** We identified 47 eligible studies. For dichotomous outcomes, we found low certainty evidence that trials without blinding of patients and healthcare providers, outcome assessors/adjudicators, and patients may slightly overestimate treatment effects. For continuous outcomes, we found low certainty evidence that trials without blinding of outcome assessors/adjudicators and patients may slightly overestimate treatment effects.

**Conclusion:** Our systematic review and meta-analysis suggests that blinding may influence trial results in select situations—albeit the findings are of low certainty and the magnitude of effect is modest. In the absence of high certainty evidence suggesting that trials with and without blinding produce similar results, investigators should be cautious about interpreting the results of trials without blinding.

## Background

Randomized trials represent the optimal design for assessing the effectiveness of interventions. Randomization ensures—more or less—that important known and unknown prognostic factors are balanced between arms such that any observed differences between trial arms can be attributed to the intervention under study. For decades, blinding—the concealment of the arm to which participants have been randomized—has been an important consideration for assessing risk of bias of randomized trials (1, 2).

Blinding of participants and healthcare providers reduces opportunity for differences in care (co-interventions)—referred to as performance bias—whereas blinding of outcome assessors/adjudicators reduces differences in the measurement and adjudication of outcomes between trial arms—called ascertainment bias (3, 4). Preconceived notions about the efficacy and safety of interventions, even subconsciously, may theoretically impact decisions about co-interventions and outcome expectancy (5, 6).

Though there are clear advantages to blinding, blinding requires significant resources, increases the operational complexity of trials, and may not always be feasible. Funding and research organizations have invested in several initiatives to streamline and simplify clinical trials, and foregoing blinding is a logical step in this process (7-9). It is unclear, however, the extent to which open-label trials may produce biased findings.

Meta-epidemiologic studies (studies that analyze the results of previous studies to address how methodological characteristics of the studies influence their results) have yielded inconsistent findings (10-15). In this systematic review and meta-analysis, we summarize evidence addressing the effects of blinding on trial results.

## Methods

We registered a protocol on Open Science Framework (https://osf.io/qenky) on June 9, 2022. We report our systematic review using the PRISMA reporting guidelines and guidelines for reporting meta-epidemiologic methods studies (16, 17).

### Search strategy

In consultation with an experienced research librarian, we searched MEDLINE, EMBASE, the Cochrane Database of Systematic Reviews, JBI EBP, and Web of Science, from inception to May 2022, for studies comparing the results of trials with and without blinding. We supplemented our search by reviewing the reference lists of eligible studies. We did not restrict the search by date or language of publication Supplement 1 presents our search strategy.

### Screening and study eligibility

Following training and calibration exercises to ensure sufficient agreement, pairs of reviewers, working independently and in duplicate, reviewed titles and abstracts of search records and subsequently the full-texts of records deemed potentially eligible at the title and abstract screening stage. Reviewers resolved discrepancies by discussion, or, when necessary, by adjudication by a third party.

We included meta-epidemiologic studies that compared the results of trials in which either patients, healthcare providers, outcome assessors/adjudicators, analysts, or manuscript writers were blinded to trials in which either of these parties were not blinded. We included studies that investigated the effects of blinding for trials addressing the effectiveness or safety of a treatment or management strategy and excluded studies that investigated the effects of blinding in diagnostic studies. We did not restrict eligibility based on the population, intervention, or outcomes or by language or date of publication.

We excluded single systematic reviews and meta-analyses that presented subgroup or sensitivity analyses addressing blinding because single reviews are unlikely to provide precise estimates and because such estimates have limited generalizability. We excluded conference abstracts and studies addressing the effects of blinding in animal studies or **in vitro** studies.

### Data extraction and risk of bias assessments

Pairs of reviewers, working independently and in duplicate, collected data and assessed the risk of bias of eligible studies.

We collected data on study characteristics (clinical condition, number of reviews/meta-analyses and trials included, type of outcome investigated), blinding characteristics (blinding status of participants, healthcare providers/investigators, outcome assessors/adjudicators, or other parties), and the results for studies with and without blinding from the least and most adjusted estimate. For studies in which multiple adjusted estimates were presented, based on previous evidence that the effects of blinding may be confounded by allocation concealment, we preferentially extracted the estimate that was adjusted for allocation concealment (10, 18).

When reported, we extracted results separately for objective and subjective outcomes. We considered outcomes objective if they were established laboratory measures and other outcomes not subject to interpretation (e.g., all-cause mortality, hospitalization) and subjective if outcomes were patient-reported or subject to interpretation from healthcare providers or investigators (e.g., adverse events).

To our knowledge, there are no risk of bias tools for meta-epidemiologic studies. We assessed the risk of bias of eligible studies using ad-hoc criteria: selection bias (association between trial characteristics and outcomes in trials selected for analysis), confounding (distortion of the effect of blinding on study results due to the correlation of the study characteristic of interest with other study characteristics that influence the results), bias in classification of trial characteristics (misclassification of blinding status), bias in measurement of outcomes (errors in extraction or reporting of results that may be related to blinding), and selective reporting bias (selective reporting of study results based on the magnitude, statistical significance, or direction of findings). To make judgements about confounding bias, we considered whether the study adjusted for allocation concealment, which has been shown to influence trial results (10, 18), and to make judgements about bias in classification of trial characteristics and measurement of outcomes, we considered the criteria used by the study to ascertain blinding status and whether trial characteristics and outcome data were collected independently and in duplicate to reduce the potential for errors (15, 19).

We rated risk of bias criteria as either low, probably low, probably high, or high risk of bias and considered results at low risk of bias overall if all domains were rated as either low or probably low risk of bias.

Reviewers resolved discrepancies by discussion or, when necessary, by third-party adjudication.

### Data synthesis and analysis

We report categorical trial characteristics as proportions and percentages and continuous characteristics as medians and interquartile ranges (IQRs). We synthesize the results of studies that compared trials with and without blinding of patients and healthcare providers/investigators (corresponding to ‘double-blind’) (20), outcome assessors/adjudicators, patients, healthcare providers/investigators, statisticians, and manuscript writers.

We performed a frequentist random-effects meta-analysis with the restricted maximum likelihood (REML) heterogeneity estimator to pool the results of studies. For studies that addressed dichotomous outcomes, we present results as ratios of odds ratios (ROR) and for studies that addressed continuous outcomes, we present differences in standardized mean differences (dSMD) with the associated confidence intervals. We recoded the data such that ROR < 1 and dSMD < 0 correspond to an exaggerated effect in trials without blinding compared to trials with blinding.

Where possible, we meta-analyzed the least and most-adjusted results from primary studies. For studies that converted dichotomous outcome measures to continuous outcome measures, or vice versa, we combined them in analyses with the measure to which they were converted. When studies reported multiple outcomes, we synthesized results for the outcome reported by the greatest number of trials.

We summarize heterogeneity using the I^2^ statistic. We considered heterogeneity of 0%–40% as potentially unimportant, 30%–60% as moderate, 50%–90% as substantial and 75%–100% as critical (21). For analyses with 10 or more studies, we assessed for publication bias using funnel plots and the Egger’s test (22).

We performed all analyses in R (version 4.03, R Foundation for Statistical Computing), using the *meta* and *metafor* packages. Open Science Framework (https://osf.io/qb23h/) presents the data and code to generate the results in this study.

Where meta-analysis is not possible, we report the range and distribution of results from primary studies (23).

### Assessment of the certainty of evidence and interpretation of study results

We use a modified GRADE approach to assess the certainty of evidence (24). While GRADE has not been developed to assess the certainty of meta-epidemiologic evidence, we anticipate that GRADE criteria are still relevant to methodologic questions and meta-epidemiologic studies.

The GRADE approach rates the evidence as either high, moderate, low, or very low certainty based on considerations of risk of bias (study limitations), inconsistency (heterogeneity in study results), indirectness (differences between the questions addressed in studies and the question of interest), publication bias (the tendency for studies with statistically significant results or positive results to be published, published faster, or published in journals with higher visibility), and imprecision (random error).

High certainty evidence indicates situations in which we are confident that the estimated effect represents the true effect, and low or very low certainty evidence indicates situations in which the estimated effect may be substantially different from the true effect.

We considered an ROR of 1.10 or 0.90 and a dSMD of 0.1 to indicate minimally important effects of blinding on trial results, which was informed by our experiences in differences in effect estimates that may influence decision-making (25, 26). Such an effect corresponds to a 10% difference in the results of blinded and unblinded trials for dichotomous outcomes and a quarter standard deviation difference for continuous outcomes.

We report our results using GRADE guidance (27), which involves describing the effect of an intervention based on the certainty of evidence (i.e., high certainty evidence the exposure is effective, moderate certainty evidence the exposure is probably effective, low certainty evidence the exposure may be effective and very low certainty evidence the effect of the exposure is unclear).

## Results

### Search results

Our search yielded a total of 6,623 unique records, of which 47 were eligible to be included (18, 28-73). Figure 1 presents the results of the search and screening.

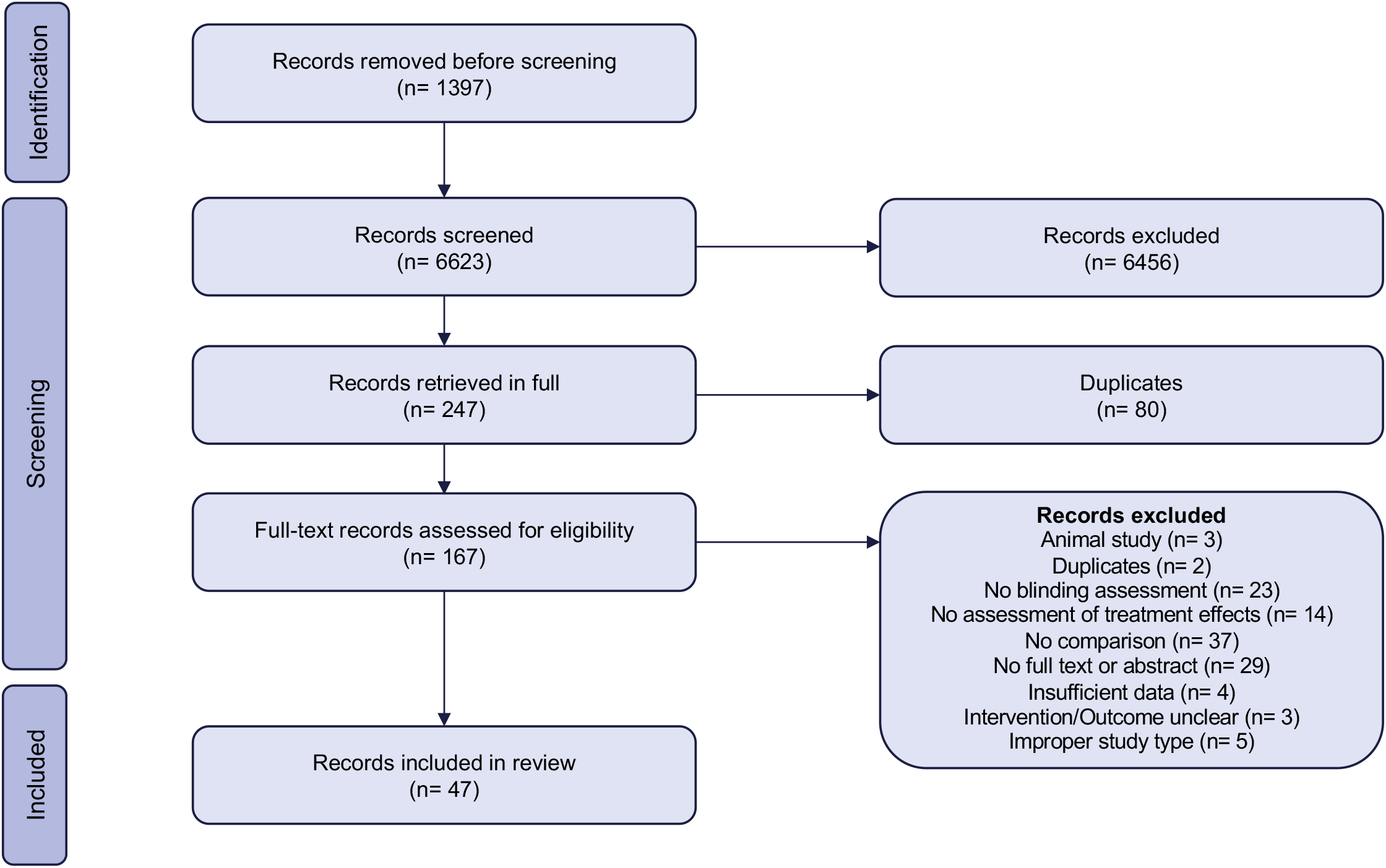

### Study characteristics

Table 1 and Supplement 2 present study characteristics. Typical studies sampled meta-analyses and compared the results of trials with and without blinding using either frequentist meta-regression or Bayesian hierarchical models (74, 75). Studies that used frequentist meta-regressions compared the results of trials with and without blinding within each meta-analysis using meta-regression and then pooled the results of coefficients corresponding to blinding across meta-analyses.

**Table 1:**
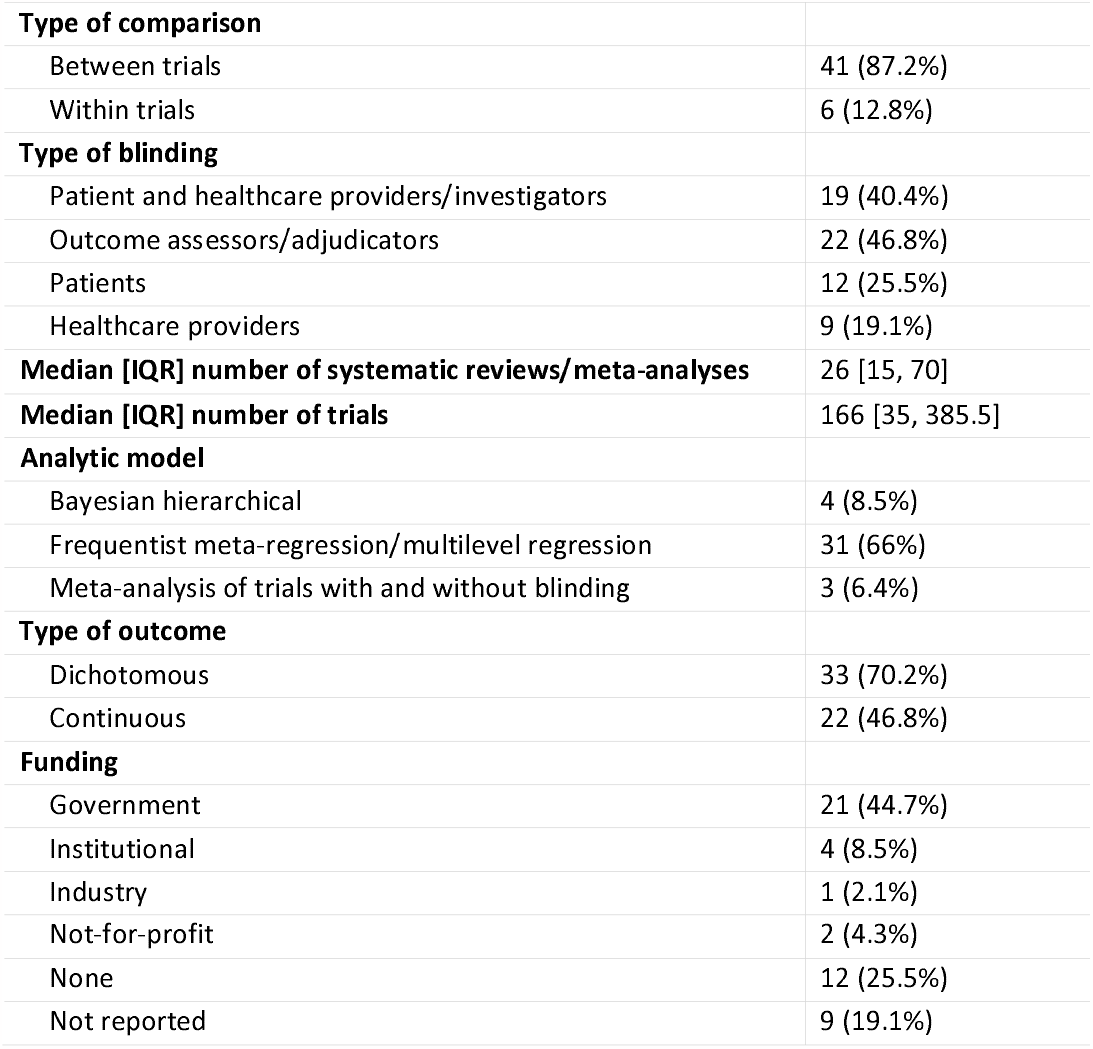
Table of study characteristics.

A minority of studies reported on studies that made within trial comparisons (37, 38, 50-52). Such studies included trials in which patients were randomized to blinded or open-label substudies or studies that performed both blinded and open-label assessment of outcomes.

Studies included a median of 26 meta-analyses and 150 trials.

### Risk of bias assessments

Figure 2 presents the risk of bias of studies that were included in the analysis investigating the effects of blinding of patients and healthcare providers/investigators and Supplement 3 presents risk of bias judgements for all studies.

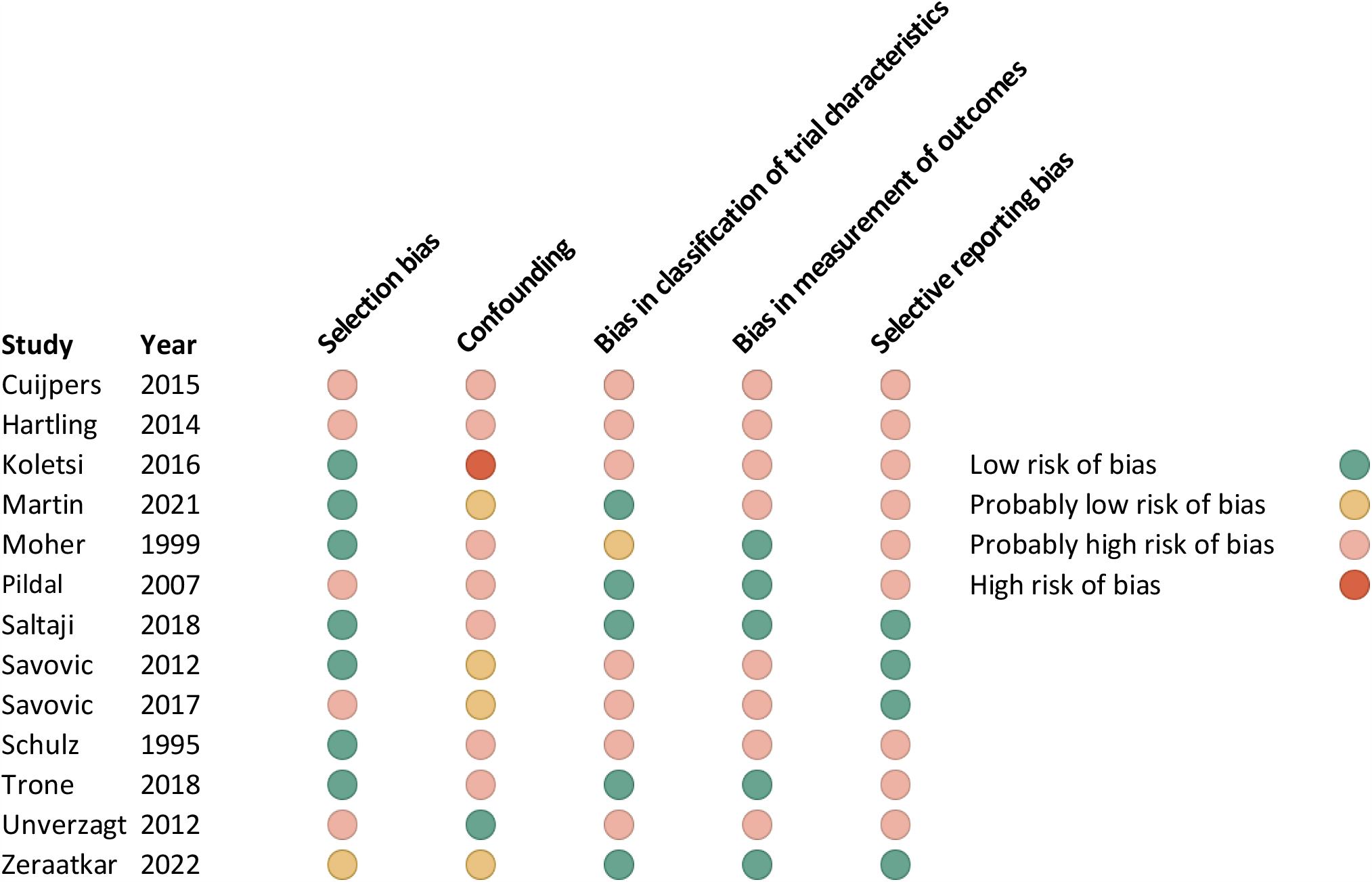

All studies were rated at high risk of bias, primarily due to confounding and selective reporting bias. Studies seldom adjusted for allocation concealment and registered or published their protocols.

### Blinding of patients and healthcare providers/investigators

Twenty-four studies addressed the effects of blinding of patients and healthcare providers/investigators on trial results (18, 28, 29, 32, 36, 39, 40, 43-46, 49, 50, 54-56, 58-60, 64-66, 69). Table 2 presents the results of our analyses.

**Table 2:**
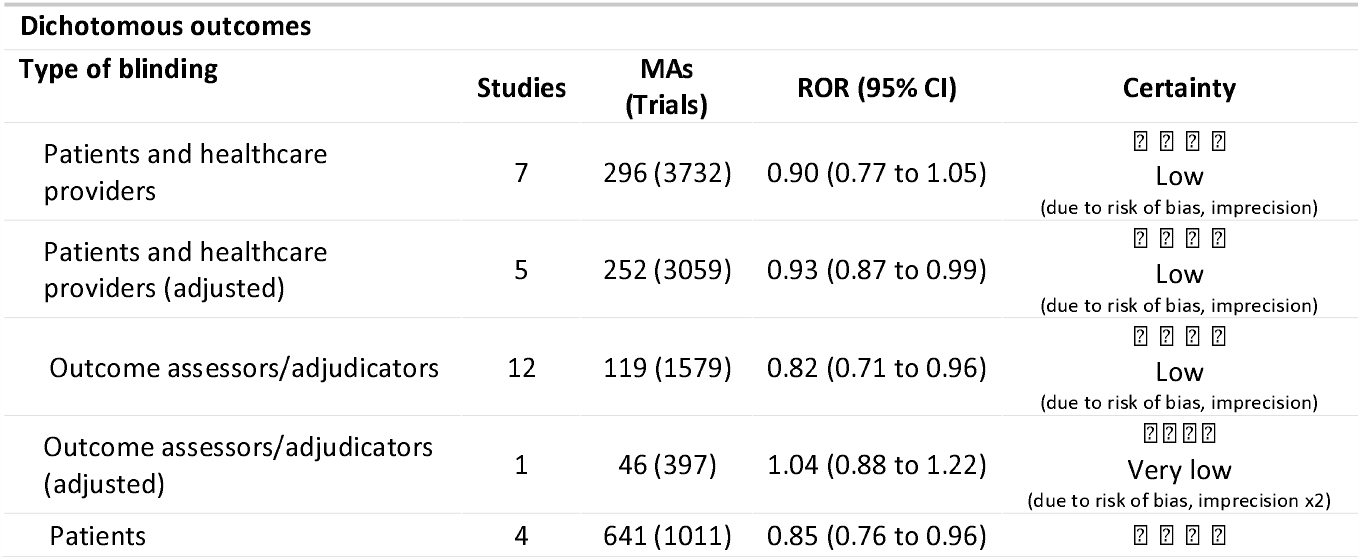
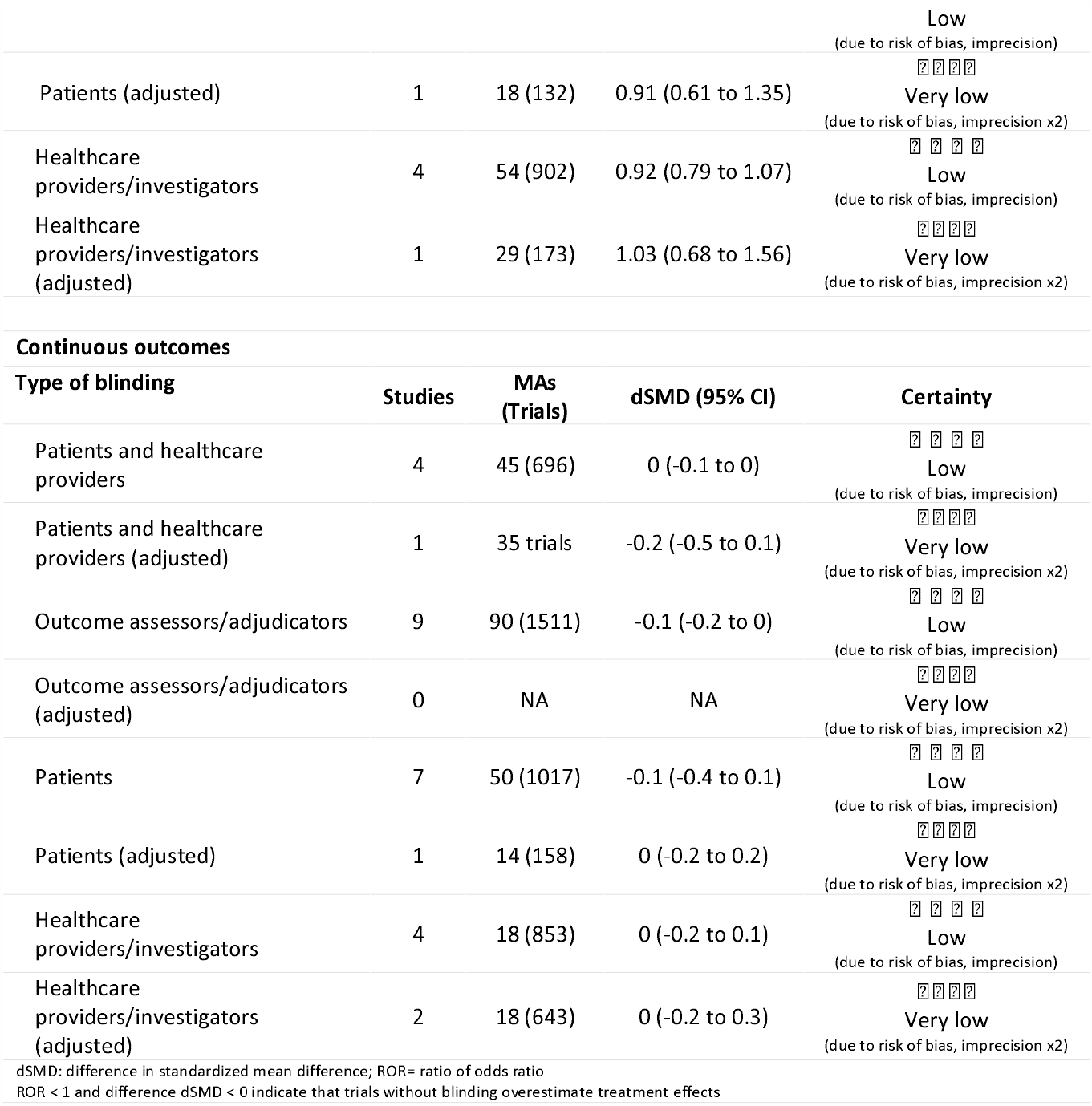
Results.

For dichotomous outcomes, we meta-analyzed seven studies, including 29546 meta-analyses and 3,732 trials, and found evidence that trials without blinding of patients and healthcare providers may overestimate dichotomous outcomes (low certainty due to risk of bias and imprecision) (Supplement 4). When we restricted our analysis to studies that reported adjusted effect estimates, we found a smaller difference between the results of trials with and without blinding.

For continuous outcomes, we meta-analyzed four studies, including 45 meta-analyses and 696 trials, and found that trials without blinding may not produce different results compared to trials with blinding (low certainty due to risk of bias and imprecision).

### Blinding of outcome assessors

Twenty-three studies addressed the effects of blinding of outcome assessors/adjudicators (30, 33-35, 37-39, 41, 42, 44, 51-53, 55, 57, 58, 61-63, 71-73).

For dichotomous outcomes, we meta-analyzed 12 studies, including 119 meta-analyses and 1,579 trials, and for continuous outcomes, we meta-analyzed 7 studies, including 78 meta-analyses and 1,426 trials, and found evidence that trials without blinding of outcome assessors may overestimate results (low certainty due to risk of bias and imprecision). We found similar results when we restricted our analyses to subjective outcomes. For dichotomous outcomes, the funnel plot showed some evidence of asymmetry but the Egger test for funnel plot asymmetry was not statistically significant (p=0.38).

### Blinding of patients

Twelve studies addressed the effects of blinding of patients (31, 33-35, 41, 44, 47, 48, 55, 62, 71, 73).

For dichotomous outcomes, we meta-analyzed four studies, including 78 meta-analyses and 1,106 trials and for continuous outcomes, we meta-analyzed seven studies, including 50 meta-analyses and 1,114 trials. We found evidence that trials without blinding of patients may overestimate treatment effects (low certainty due to risk of bias and imprecision). The results were similar when we restricted the analysis to subjective outcomes.

### Blinding of healthcare providers/investigators

Nine studies addressed the effects of blinding of healthcare providers (33-35, 41, 44, 55, 63, 71, 73).

We meta-analyzed four studies reporting on dichotomous outcomes, including 54 meta-analyses and 902 trials, and four studies reporting on continuous outcomes, including 18 meta-analyses and 859 trials. For both dichotomous and continuous outcomes, we found that trials without blinding of healthcare providers may not overestimate treatment effects (low certainty due to risk of bias and imprecision).

### Statisticians and manuscript writers

Two studies addressed the effects of blinding of statisticians, one of which did not find sufficient data, and none of the studies addressed the effects of blinding of manuscript writers (34, 55).

The one study with data addressing the effects of blinding of statisticians found that blinding of statisticians may have no effect on trial results but that the results are very uncertain (very low certainty due to risk of bias, imprecision, and indirectness).

## Discussion

### Main findings

Our systematic review presents evidence that trials without blinding may overestimate treatment effects—albeit the findings are of low certainty and the magnitude of effect is modest.

We found inconsistencies in the effects of blinding. While we found evidence, for example, that trials without blinding of patients and healthcare providers may overestimate dichotomous outcomes, we did not find evidence for such an effect for continuous outcomes. Similarly, we also found inconsistencies across studies. The MetaBLIND study, for example, did not find evidence that trials with and without blinding produce different results—contrary to several other studies of similar magnitude included in our review (40, 41, 59, 69). Further, studies that compared the effects of blinding within rather than across trials showed blinding to have larger effects (38, 50-52).

Theoretical considerations about blinding may explain some of the observed inconsistencies. For example, we anticipate several factors that may vary based on the question being investigated and that may influence the effect of blinding: investigators’ and patients’ preconceived notions about the effectiveness of the intervention; whether there are co-interventions that may influence the outcome of interest; the magnitude of effect of the co-interventions on the outcome of interest; the potential for imbalances in the administration of co-interventions; and the degree of interpretation involved in assessment of outcomes (18, 76).

An open-label trial addressing a condition for which there are no effective co-interventions or a trial in which healthcare providers and patients either do not have access to co-interventions or in which the administration of co-interventions is strictly regimented across arms is unlikely to be at risk of performance bias. Similarly, an open-label trial addressing outcomes that are objective and not subject to interpretation is unlikely to be at risk of ascertainment bias.

In the absence of high certainty evidence suggesting that trials with and without blinding produce similar results, investigators should still be cautious about open-label trials.

### Strengths and limitations

The strengths of our systematic review include *a priori* methods, a systematic search for eligible studies, duplicate screening and data extraction, and assessment of the role of blinding across a range of clinical areas. Our review improves on previous reviews by performing an assessment of risk of bias and the certainty of evidence and including several recently published studies not included in previous reviews (10, 15, 18, 19, 33, 40, 44).

Our results are limited by the risk of bias of primary studies. Nearly all studies were rated at high risk of bias, primarily due to potential for confounding and selective reporting bias. There is also potential for misclassification of blinding status. A substantial proportion of trials, for example, did not explicitly report the blinding status for all relevant parties (77). When trials did not explicitly report blinding status, studies either used validated guidance to ascertain blinding status or grouped unblinded studies with studies with unclear blinding (78). Even when blinding status was reported, the reporting may not have been accurate (79). Several studies used the risk of bias judgements from meta-analyses as a surrogate for blinding status, which may also contain errors or may reflect reviewers’ judgements regarding the potential impact of blinding or lack thereof on results rather than whether the trial was blinded. This potential for misclassification means that the role of blinding may be more important than our estimates suggest.

Our results are further limited by the potential inconsistency in the direction of the effect of blinding. While we anticipate that trials without blinding overestimate the beneficial effects of treatments, it is possible that the direction of bias caused by lack of blinding may not be consistent. That is, in certain circumstances open-label trials may overestimate effects and in other circumstances they may underestimate effects.

All of our analyses produced results that were imprecise. Further, our estimated effects for blinding were generally small and evidence users may consider such effects too small to influence decision-making, though most medical treatments show modest effects and hence even small biases may impact decision-making (80, 81).

Trials addressing questions that are sensitive to performance bias and ascertainment bias may be more likely to be blinded, thus reducing observed differences between trials with and without blinding.

There may be overlap between trials and meta-analyses included in meta-epidemiologic studies. An inspection of the search strategies and clinical areas addressed by the included studies, however, suggests that this overlap is likely minimal. To reduce the potential for double-counting, when meta-epidemiologic studies reported more than one estimate that would be eligible to be included in each analysis, we only included the estimate from the analysis that included the greatest number of trials.

While we tried to pool studies addressing the effects of blinding different individuals together (e.g., patients, healthcare providers), the ways in which types of blinding were defined and operationalized across studies may have been slightly different.

It is possible that blinding may be compromised during a trial (e.g., due to obvious adverse events, poorly matched placebos, or large therapeutic effects), which may have attenuated the difference between trials with and without blinding.

### Implications

The results of our systematic review have implications for evidence users who are appraising and applying evidence from open-label trials, for clinical trialists considering the potential benefits and burdens of implementing blinding in their clinical trials, and for global efforts to streamline and simplify clinical trial methods (7-9).

Results from the MetaBLIND study encouraged a critical evaluation of the importance of blinding in clinical trials (41). The study, for example, led to a revision of the Cochrane-endorsed risk of bias tool (RoB 2.0) to include a more nuanced approach for assessing bias due to lack of blinding (76). Results from MetaBLIND, however, were imprecise and were consistent with lack of blinding resulting in both an over- and underestimation of treatment effects. Further, there is no evidence whether the new approach to risk of bias assessments, proposed by the RoB 2.0 tool, that places less emphasis on blinding is better correlated with trial results.

The imprecision in results across meta-epidemiologic studies may be driven by heterogeneity in the effects of blinding across trials. That is, in some situations trials without blinding may overestimate treatment effects. Our findings suggest that evidence users should be cognizant of the ways in which lack of blinding may bias trial results, that clinical trialists should be judicious in their decisions to forego blinding, and that evidence users should be cognizant of the ways in which lack of blinding may bias trial results.

Evidence users should be mindful that the inability to identify evidence of when and how blinding affects trial results does not mean that open-label designs are unbiased. In fact, our review shows evidence that trials without blinding may overestimate treatment effects.

The subject of blinding invites comparisons to judgements about the quality of evidence from non-randomized studies. Despite evidence that randomized trials and non-randomized studies produce consistent results in many situations (82-84), we typically consider non-randomized studies to produce lower certainty evidence compared to randomized trials because evidence users are unable to confidently distinguish between situations in which non-randomized studies produce trustworthy or untrustworthy results. Uncertainty about the effects of blinding may analogously demand always or almost always rating open-label studies at high risk of bias.

We show that we need higher certainty evidence to determine when and how blinding affects trial results. Initiatives such as Studies Within a Trial (SWATs)—research studies embedded within a host trial to assess alternative ways of delivering or organizing a particular trial process—can help address this need (85). While incorporating SWATs within trials to study how blinding affects results may increase the costs of trials, such initiatives may prove cost-effective in the long term by simplifying and streamlining future trials without compromising the trustworthiness of results.

## Conclusion

Our systematic review and meta-analysis presents evidence that trials without blinding may overestimate treatment effects—albeit the findings are of low certainty and the magnitude of effect is modest. In the absence of high certainty evidence suggesting that trials with and without blinding produce similar results, investigators should be cautious about interpreting the results of trials without blinding.

## Data Availability

All data produced in the present study are available upon reasonable request to the authors

## Acknowledgements

None.

