## Supplementary material for "The impact of blinding on trial results: A systematic review and meta-analysis"

Dena Zeraatkar

### Supplement 1: Search Strategy

| Ovid MEDLINE(R) ALL <1946 to May 27, 2022> | 1144 |
| --- | --- |
| Embase 1974 to 2022 May 27 | 5726 |
| Cochrane Reviews | 14 |
| JBI EBP Database Current to May 18, 2022 | 0 |
| Web of Science Core Collection – Science Citation Index Expanded 1976 - present | 1136 |

Database: Ovid MEDLINE(R) ALL <1946 to May 27, 2022>

Search Strategy:

--------------------------------------------------------------------------------

1	Meta-Analysis as Topic/ or Meta-Analysis/ (181273)

2	(meta-analy* or metaanaly* or pooled analy?s).tw,pt,kw. (260393)

3	exp Review Literature as Topic/ or Systematic Review/ (216029)

4	(systematic adj (review*1 or overview*1)).tw,pt,kw. (244596)

5	(Meta-Epidemiologic* or Metaepidemiologic*).mp. (310)

6	1 or 2 or 3 or 4 or 5 (413481)

7	Randomized Controlled Trials as Topic/ or randomized controlled trial/ or Random Allocation/ or clinical trials as topic/ (979183)

8	(((clinical or control*) adj2 trial*) or random*).tw,kf. (1637883)

9	7 or 8 (1969824)

10	*single-blind method/ or *Double-Blind Method/ (861)

11	(Blind* or blinding or nonblind* or non-blinding).ti. (72324)

12	bias.ti. (22253)

13	10 or 11 or 12 (94607)

14	6 and 9 and 13 (1144)

***************************

Database: Embase <1974 to 2022 May 27>

Search Strategy:

--------------------------------------------------------------------------------

1	"meta analysis (topic)"/ or exp Meta Analysis/ (292882)

2	(meta-analy* or metaanaly* or pooled analy?s).tw,pt,kw. (305126)

3	"systematic review (topic)"/ or exp "systematic review"/ (371037)

4	Randomized Controlled Trials as Topic/ or randomized controlled trial/ or randomization/ or Double Blind Procedure/ or Single Blind Procedure/ or exp Clinical Trial/ or exp controlled clinical trial/ (1941080)

5	(((clinical or control*) adj2 trial*) or random*).tw,kw. (2249130)

6	single blind procedure/ or double blind procedure/ (239350)

7	(Blind* or blinding or nonblind* or non-blinding).ti. (90559)

8	bias.ti. (24224)

9	1 or 2 or 3 (574838)

10	4 or 5 (3198006)

11	6 or 7 or 8 (303123)

12	9 and 10 and 11 (5726)

***************************

Database: Web of Science

Search Strategy:

--------------------------------------------------------------------------------

1.	TS = (Meta-analys?s)

2.	TI = (meta-analy* or metaanaly* or pooled analy?s)

3.	AB = (meta-analy* or metaanaly* or pooled analy?s)

4.	TI = (systematic NEAR/1 (review* or overview*))

5.	AB = (systematic NEAR/1 (review* or overview*))

6.	TI = (Meta-Epidemiologic* or Metaepidemiologic*)

7.	AB = (Meta-Epidemiologic* or Metaepidemiologic*)

8.	TS = ((randomi?ed NEAR/2 trial) or (clinical NEAR/1 trial))

9.	TI = (((clinical or control*) NEAR/2 trial*) or random*)

10.	AB = (((clinical or control*) NEAR/2 trial*) or random*)

11.	#10 OR #9 OR #8

12.	TI = (Blind* or blinding or nonblind* or non-blinding or bias)

13.	TS = ((single-blind NEAR/1 method) or (double-blind NEAR/1 method))

14.

#12 OR #13

15.	TS = ((systematic NEAR/1 review) or (literature NEAR/1 review))

16.	#1 OR #2 OR #3 OR #4 OR #5 OR #6 OR #7

17.	#14 AND #11 AND #16

Database: JBI EBP Database <Current to May 18, 2022>

Search Strategy:

--------------------------------------------------------------------------------

1	exp meta-analysis/ (14)

2	(meta-analy* or metaanaly* or pooled analy?s).ti,ab,pt,kw. (469)

3	exp "systematic review"/ (141)

4	(systematic adj (review*1 or overview*1)).ti,ab,pt,kw. (1935)

5	(Meta-Epidemiologic* or Metaepidemiologic*).mp. (2)

6	exp clinical trial/ or exp randomized controlled trial/ (1)

7	(((clinical or control*) adj2 trial*) or random*).ti,ab,pt,kw. (449)

8	1 or 2 or 3 or 4 or 5 (1956)

9	6 or 7 (450)

10	(Blind* or blinding or nonblind* or non-blinding).ti. (0)

11	bias.ti. (2)

12	10 or 11 (2)

13. 8 and 9 and 12 (0)

***************************

Database: Cochrane Reviews <Current to May 30, 2022>

Search Strategy:

--------------------------------------------------------------------------------

ID	Search	Hits

#1	MeSH descriptor: [Meta-Analysis as Topic] explode all trees	375

#2	MeSH descriptor: [Systematic Review] explode all trees	0

#3	(Meta-Epidemiologic* or Metaepidemiologic*):ti,ab,kw	23

#4	(meta-analy* or metaanaly* or pooled analysis or pooled analyses):ti,ab,kw	30095

#5	(systematic NEAR/1 (review* or overview*)):ti,ab,kw	12558

#6	#1 or #2 or #3 or #4 or #5	36226

#7	MeSH descriptor: [Randomized Controlled Trials as Topic] explode all trees	15169

#8	MeSH descriptor: [Randomized Controlled Trial] explode all trees	119

#9	(((clinical or control*) NEAR/2 trial*) or random*):ti,ab,kw	1224912

#10	MeSH descriptor: [Single-Blind Method] explode all trees	22897

#11	MeSH descriptor: [Double-Blind Method] explode all trees	146886

#12	(Blind* or blinding or nonblind* or non-blinding):ti	101257

#13	bias:ti	1597

#14	#7 or #8 or #9	1224961

#15	#10 or #11 or #12 or #13	234503

#16	#6 and #14 and #15	2285

Limit 16 to Cochrane Reviews – 14

### Supplement 2: Table of study characteristics

| Supplement 2: Table of study characteristics | |  |  |  |  |  |  |  |
| --- | --- | --- | --- | --- | --- | --- | --- | --- |
| Study | **Search Strategy** | **Clinical area** | **Number of MAs** | **Number of trials** | **Type of outcome** | **Type(s) of blinding** | **Within vs. Between trials** | **Analytic model** |
| Amer, 2021 | Cochrane Library, inception to 2015 | open and laparoscopic abdominal surgical procedures | NA | 316 | dichotomous, continuous | outcome assessors/adjudicators, patients, healthcare providers/investigators | between trials | Frequentist meta-regression/multilevel regression |
| Anthon, 2018 | CDSR, inception to 2017 | critical care | NR | 361 | dichotomous | patient and healthcare providers/investigators | between trials | Meta-analysis with subgroup analyses |
| Armijo-Olivo, 2016 | CDSR, 2005 to 2011 | physical therapy | 43 | 393 | continuous | outcome assessors/adjudicators, patients, healthcare providers/investigators, statisticians | between trials | Frequentist meta-regression/multilevel regression |
| Baiardo Redaelli, 2018 | PubMed, 2000 to 2005 | surgical interventions in critical care | NA | 119 | dichotomous | patient and healthcare providers/investigators | between trials | Meta-analysis of results of blinded versus unblinded trials |
| Balk, 2002 | MEDLINE, inception to 2000, CDSR, 2000, issue 4 | cardiovascular disease, infectious disease, pediatrics, surgery | 26 | 276 | dichotomous | patient and healthcare providers/investigators, outcome assessors/adjudicators, patients, healthcare providers/investigators, statisticians | between trials | Bayesian hierarchical |
| Bialy, 2014 | CDSR, 2009, issue 2 | neonatal | 25 | 208 | dichotomous | outcome assessors/adjudicators, healthcare providers/investigators | between trials | Frequentist meta-regression/multilevel regression |
| Bolvig, 2018 | CDSR, NR | osteoarthritis | 20 | 126 | continuous | outcome assessors/adjudicators, patients, healthcare providers/investigators | between trials | Frequentist meta-regression/multilevel regression |
| Braithwaite, 2018 | MEDLINE, EMBASE, AMED, Scopus, CINAHL, PEDro, Cochrane Library, inception to 2016 | dry needling and pain | NA | 25 | continuous | patients | between trials | Frequentist meta-regression/multilevel regression |
| Chaimani, 2013 | PubMed, inceptiont to 2011 | no restrictions | 20 | 358 | dichotomous | outcome assessors/adjudicators, patients | between trials | network meta-epidemiological model |
| Contopoulos-Ioannidis, 2005 | Mental Health Library, 2002, issue 1 | mental health | 16 | 133 | dichotomous, continuous | patient and healthcare providers/investigators | between trials | meta-analysis of results of blinded versus unblinded trials |
| Cuijpers, 2015 | PubMed, PsycInfo, EMBASE, Cochrane Central Register of Controlled Trials, 2014 | depression | NA | 35 | continuous | patient and healthcare providers/investigators | between trials | Frequentist meta-regression/multilevel regression |
| Dello Russo, 2020 | clinicaltrials.gov, clinicaltrialsregister.eu, 2020 | oncology | NA | 20 | continuous | outcome assessors/adjudicators | within trials | Mean across trials |
| Deschartres, 2014 | CDSR, 2008 to 2010, 2011, to 2013, 10 journals with the highest impact factor for each medical specialty | no restrictions | 163 | 1240 | dichotomous | objective outcomes or blinding of participants, healthcare providers, and outcome assessors | between trials | Frequentist meta-regression/multilevel regression |
| Diakou, 2016 | Cochrane Register of Controlled Trials, PubMed, EMBASE, PsycINFO, CINAHL, Google Scholar, inception to 2015 | no restrictions | NA | 2 | dichotomous | outcome assessors/adjudicators | within trials | Frequentist meta-regression/multilevel regression |
| Egger, 2003 | American Journal of Cardiology, Annals of Internal Medicine, BMJ, Cancer, Circulation, JAMA, Lancet, Obstetrics and Gynecology, 1994 to 1998 | no restrictions | 122 | 399 | dichotomous | patient and healthcare providers/investigators | between trials | Frequentist meta-regression/multilevel regression |
| Fenwick, 2008 | CDSR, inception to 2007 | periodontology | 5 | 35 | continuous | outcome assessors/adjudicators | between trials | Frequentist meta-regression/multilevel regression |
| Feys, 2014 | PubMed, EMBASE, Cochrane Central Register of Controlled Trials, clinicaltrials.gov, Food and Drug Administration clinical reviews, inception to 2012 | PDE-5 inhibitors for erectile dysfunction | NA | 110 | continuous | patients | between trials | meta-analysis with subgroup analyses |
| Hartling, 2014 | CDSR, 2009, issue 2 | child health | 17 | 287 | dichotomous, continuous | patient and healthcare providers/investigators, outcome assessors/adjudicators | between trials | Frequentist meta-regression/multilevel regression |
| Hempel, 2012 | prior AHRQ-funded systematic reviews (dataset 2) | no restrictions | NR | 630 | dichotomous | outcome assessors/adjudicators, patients, healthcare providers/investigators | between trials | Frequentist meta-regression/multilevel regression |
| Howard, 2016 | MEDLINE, EMBASE, 2009 to 2014 | hypertension | NA | 148 | continuous | patient and healthcare providers/investigators | between trials | meta-analysis of results of blinded versus unblinded trials |
| Hrobjartsson, 2017 | PubMed, Embase, PsycINFO, CINAHL, Cochrane Central Register of Controlled Trials, High Wire Press, Google Scholar, 2013 | no restrictions | NA | 18 | continuous | outcome assessors/adjudicators | within trials | Meta-analysis of within-trial comparisons |
| Hrobjartsson (2012), 2012 | PubMed, EMBASE, PsycINFO, CINAHL, Cochrane Central Register of Controlled Trials, HighWire Press, Google Scholar, Food and Drug Administration website, inception to 2010 | no restrictions | NA | 21 | dichotomous | outcome assessors/adjudicators | within trials | Meta-analysis of within-trial comparisons |
| Hrobjartsson 2013 | PubMed, EMBASE, PsycINFO, CINAHL, Cochrane Central Register of Controlled Trials, HighWire Press, Google Scholar, Food and Drug Administration website, inception to 2010 | no restrictions | NA | 16 | continuous | outcome assessors/adjudicators | within trials | Meta-analysis of within-trial comparisons |
| Hrobjartsson, 2014 | MEDLINE, EMBASE, Cochrane methodology register, inception to 2013 | complementary/alternative medicine | NA | 12 | dichotomous, continuous | patient and healthcare providers/investigators | within trials | Meta-analysis of within-trial comparisons |
| Ioannidis, 1997 | MEDLINE, inception to 1996 | HIV | NA | 15 | dichotomous | patient and healthcare providers/investigators | between trials | meta-analysis of results of blinded versus unblinded trials |
| Juni, 1999 | previously published meta-analysis | low molecular weight heparin vs. standard heparin | NA | 17 | dichotomous | outcome assessors/adjudicators | between trials | Frequentist meta-regression/multilevel regression |
| Kjaergard, 2001 | Cochrane Library, MEDLINE, NR | no restrictions | 14 | 190 | dichotomous | patient and healthcare providers/investigators | between trials | Frequentist meta-regression/multilevel regression |
| Koletsi, 2016 | 50 most recent issues of the American Journal of Orthodontics and Dentofacial Orthopedics (AJODO), the Angle Orthodontist (Angle), the European Journal of Orthodontics (EJO), the Journal of Orthodontics (JO), 2013 | orthodontics | NA | 101 | dichotomous, continuous | patient and healthcare providers/investigators, outcome assessors/adjudicators | between trials | Frequentist meta-regression/multilevel regression |
| Lega, 2013 | MEDLINE, EMBASE, inception to 2012 | new oral anticoagulants for non-valvular atrial fibrillation | NA | 13 | dichotomous | patient and healthcare providers/investigators, outcome assessors/adjudicators | between trials | Frequentist meta-regression/multilevel regression |
| Liu, 2011 | systematic review by the Bone, Joint, and Muscle Trauma Group of Cochrane | progressive resistance training in older adults | 1 | 73 | continuous | outcome assessors/adjudicators | between trials | Frequentist meta-regression/multilevel regression |
| Martin, 2021 | five highest impact medical journals, six highest impact critical care journals, CDSR, 2009 to 2019 | critical care | 36 | 467 | dichotomous | patient and healthcare providers/investigators | between trials | Frequentist meta-regression/multilevel regression |
| Moher, 1999 | MEDLINE, EMBASE, inception to 1995, CDSR, 1995, issue 2 | no restrictions | 11 | 127 | dichotomous | patient and healthcare providers/investigators | between trials | Frequentist meta-regression/multilevel regression |
| Moustgaard, 2020 | CDSR, 2013, issue 2 | no restrictions | 142 | 1153 | dichotomous, continuous | outcome assessors/adjudicators, patients, healthcare providers/investigators | between trials | Bayesian hierarchical |
| Nuesch, 2009 | Cochrane Library, MEDLINE, EMBASE, CINAHL, inception to 2007 | osteoarthritis | 10 | 122 | continuous | patients | between trials | Frequentist meta-regression/multilevel regression |
| Pildal, 2007 | PubMed, 2001 to 2002, CDSR, 2003, issue 2 | no restrictions | 70 | 499 | dichotomous | patient and healthcare providers/investigators | between trials | Frequentist meta-regression/multilevel regression |
| Poolman, 2007 | The Journal of Bone and Joint Surgery, 2003 to 2004 | orthopedics | NA | 32 | dichotomous, continuous | outcome assessors/adjudicators | between trials | Frequentist meta-regression/multilevel regression |
| Probst, 2019 | Cochrane Central Register of Controlled Trials, MEDLINE, Web of Science, 2015 | general and abdominal surgery | NA | 378 | NR | patient and healthcare providers/investigators | between trials | Frequentist meta-regression/multilevel regression |
| Saltaji, 2018 | PubMed, MEDLINE, EMBASE, ISI Web of Science, CDSR, Health STAR, American Dental Association (ADA)–Evidence based Dentistry database, inception to 2014 | oral health | 64 | 540 | continuous | patient and healthcare providers/investigators, outcome assessors/adjudicators, patients, healthcare providers/investigators | between trials | Frequentist meta-regression/multilevel regression |
| Savovic, 2012 | Data from Egger 2003, Schulz 1995, Balk 2002, Pildal 2007, Kjaergard 2007, Siersma 2007 | no restrictions | 104 | 590 | dichotomous | patient and healthcare providers/investigators | between trials | Bayesian hierarchical |
| Savovic, 2017 | CDSR, 2011, issue 4 | no restrictions | 144 | 1678 | dichotomous | patient and healthcare providers/investigators | between trials | Bayesian hierarchical |
| Schulz, 1995 | Cochrane Pregnancy and Childbirth Database, 1993 | pregnancy and childbirth | 33 | 250 | dichotomous | patient and healthcare providers/investigators | between trials | Frequentist meta-regression/multilevel regression |
| Siersma, 2007 | CDSR, NR | no restrictions | 48 | 523 | dichotomous | double-blinding or blinding of outcome assessor | between trials | Frequentist meta-regression/multilevel regression |
| Trone, 2018 | MEDLINE, Cochrane, clinicaltrials.gov, 2003 to 2016 | antiangiogenic therapies and vascular adverse drug events | NA | 166 | dichotomous | patient and healthcare providers/investigators | between trials | Frequentist meta-regression/multilevel regression |
| Unverzagt, 2013 | CDSR, issue 1, 2011 | sepsis and shock | 12 | 82 | dichotomous | patient and healthcare providers/investigators | between trials | Frequentist meta-regression/multilevel regression |
| van Tulder, 2009 | CDSR, 2005, issue 3 | lower back pain | 15 | 216 | dichotomous, continuous | outcome assessors/adjudicators, patients, healthcare providers/investigators | between trials | Frequentist meta-regression/multilevel regression |
| Wood, 2008 | Data from Schulz 1995, Kjaergard 2001, Egger, 2003 | no restrictions | 76 | 746 | dichotomous | double-blinding or blinding of outcome assessor | between trials | Frequentist meta-regression/multilevel regression |
| Zeraatkar, 2022 | Living COVID-19 Systematic Reviews | COVID-19 | NA | 352 | dichotomous, continuous | patient and healthcare providers/investigators | between trials | Frequentist meta-regression/multilevel regression |
| CDSR: Cochrane Database of Systematic Reviews; MA: meta-analysis; NR: not reported | | | | | | | | |

### **Supplement 3: Risk of bias assessments for studies contributing to meta-analyses**

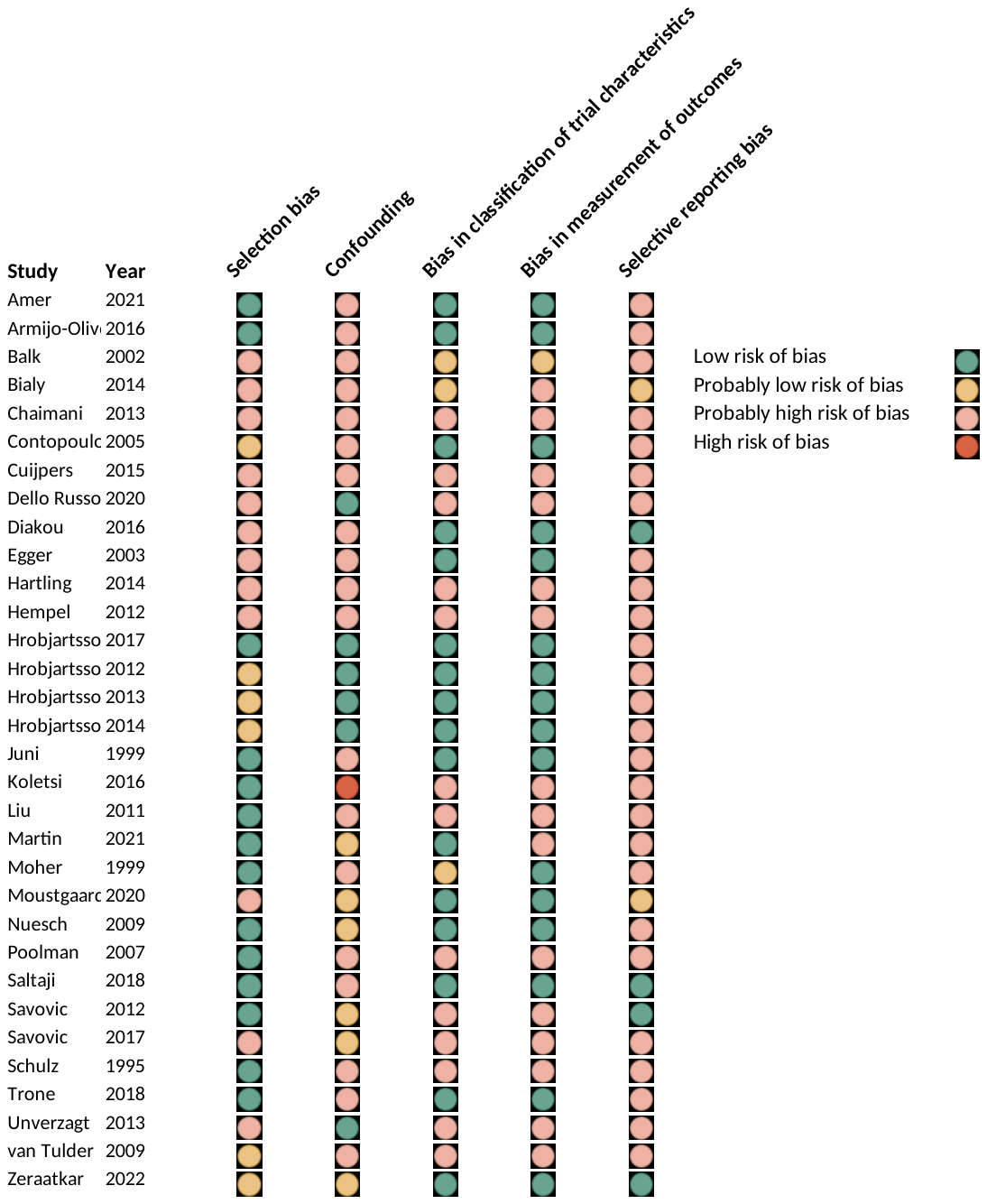

### Supplement 4: Forest plots

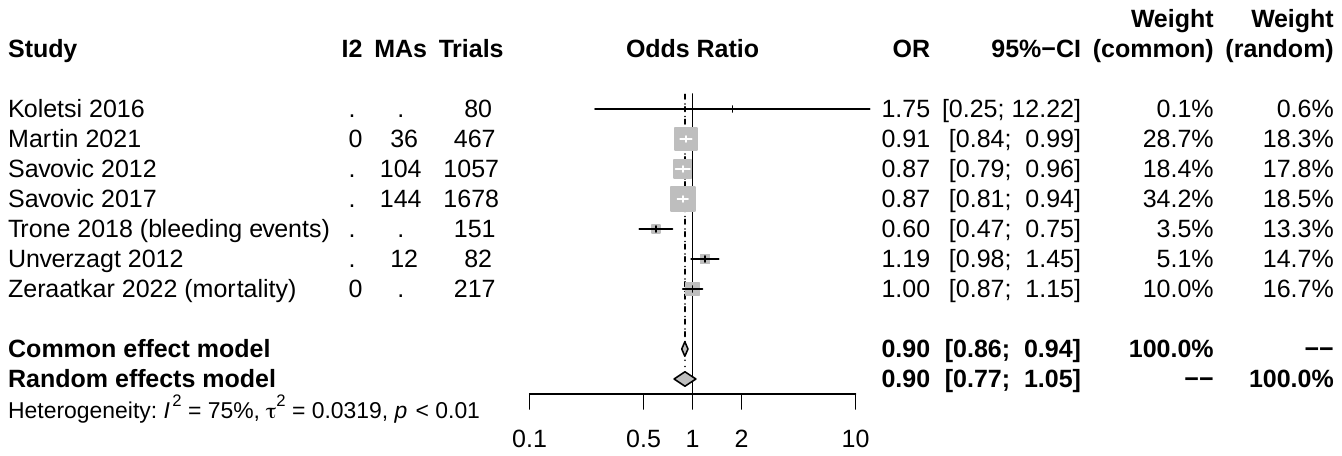

#### Supplemental figure 4.1. Forest plot of patient and healthcare provider blinding for dichotomous outcomes in double-blind vs open-label studies.

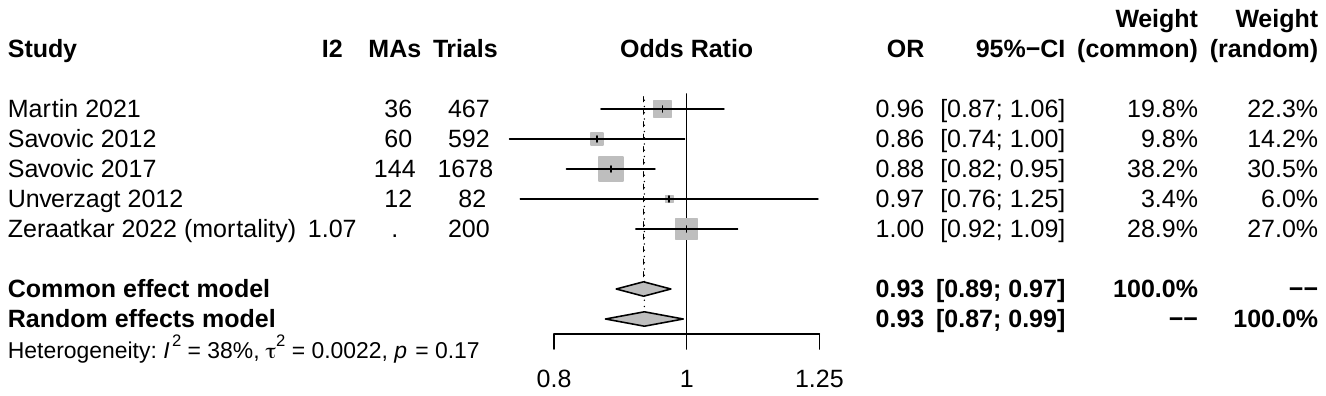
Supplemental figure 4.2. Forest plot of patient and healthcare provider blinding (adjusted) for dichotomous outcomes in double-blind vs open-label studies.

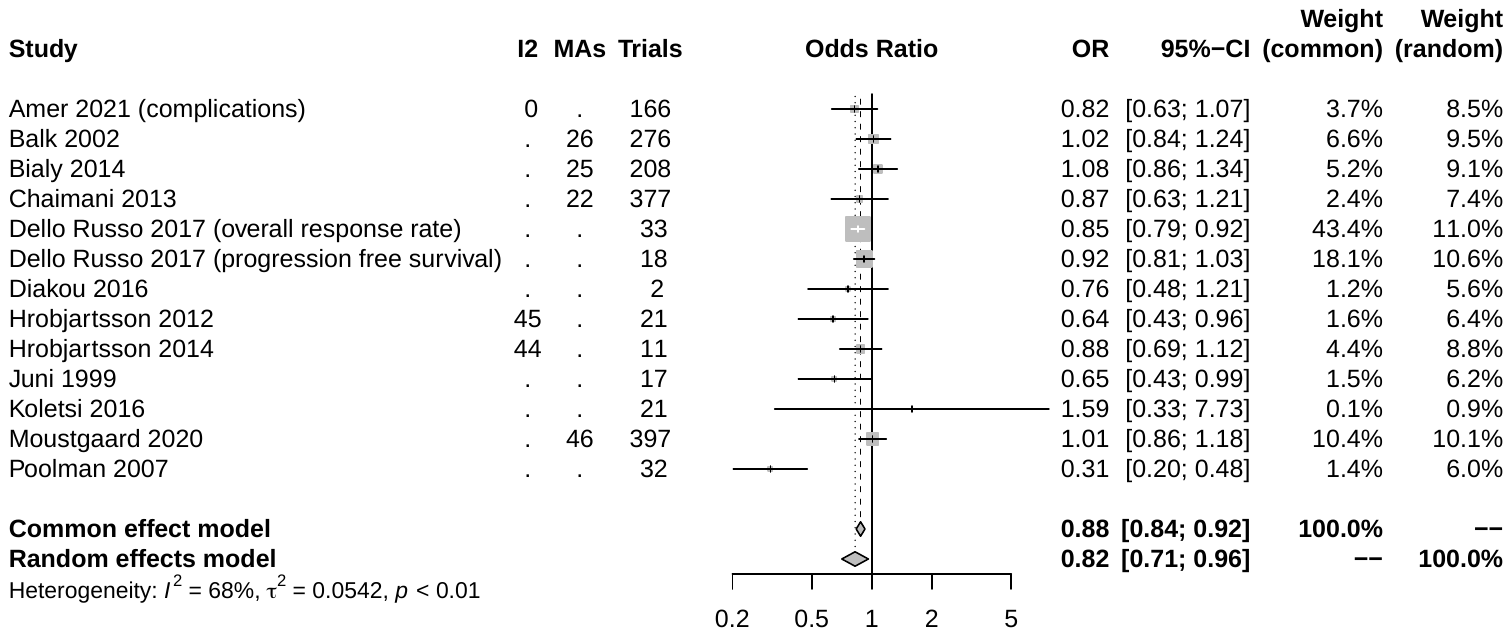

Supplemental figure 4.3. Forest plot of outcome assessor/adjudicator blinding for dichotomous outcomes in double-blind vs open-label studies.
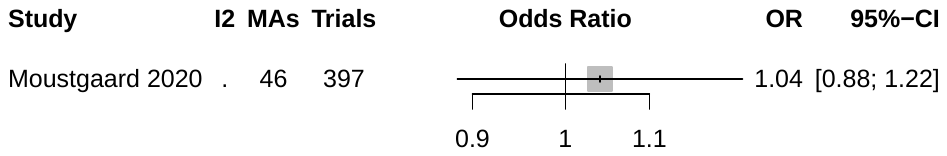

#### Supplemental figure 4.4. Forest plot of outcome assessor/adjudicator blinding (adjusted) for dichotomous outcomes in double-blind vs open-label studies.

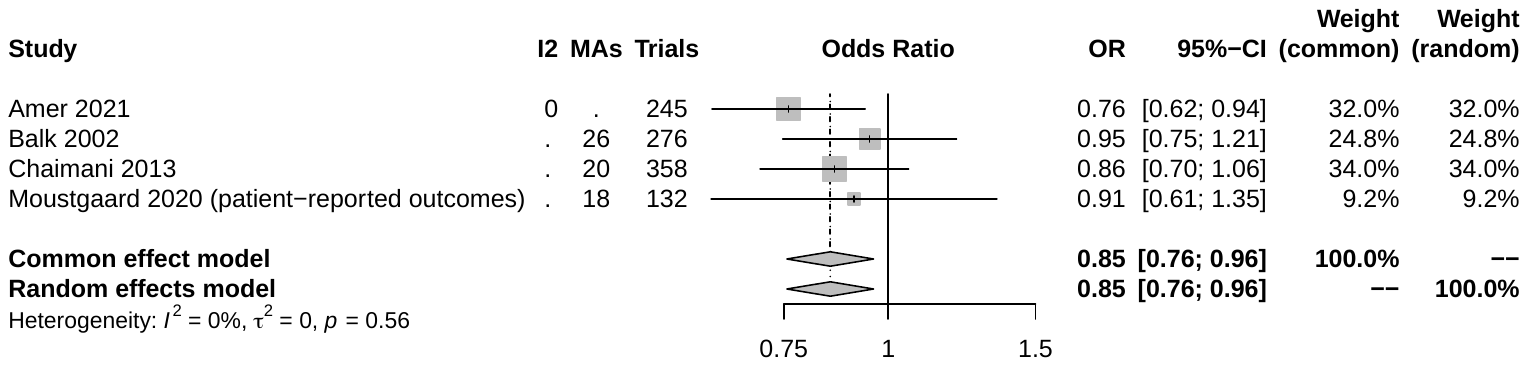

#### Supplemental figure 4.5. Forest plot of patient blinding for dichotomous outcomes in double-blind vs open-label studies.

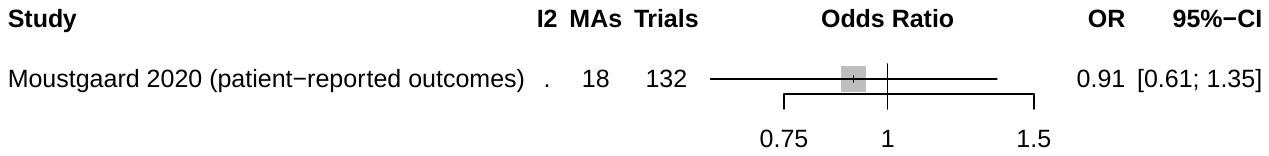

#### Supplemental figure 4.6. Forest plot of patient blinding (adjusted) for dichotomous outcomes in double-blind vs open-label studies.

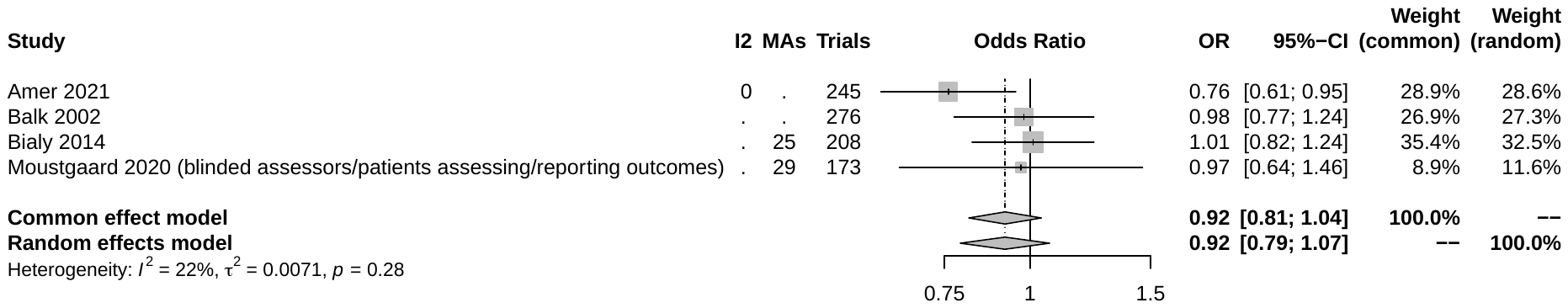

#### Supplemental figure 4.7. Forest plot of healthcare provider blinding for dichotomous outcomes in double-blind vs open-label studies.

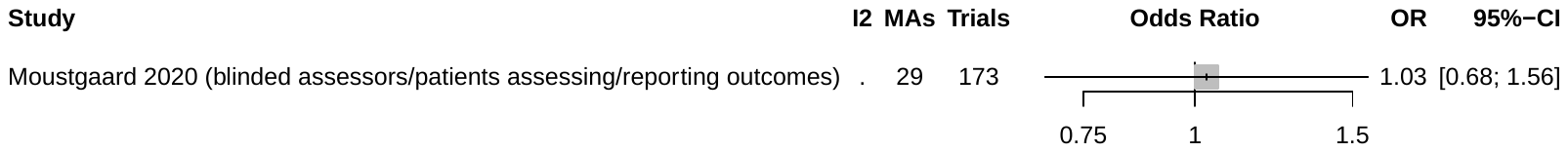

#### Supplemental figure 4.8. Forest plot of healthcare provider blinding (adjusted) for dichotomous outcomes in double-blind vs open-label studies.

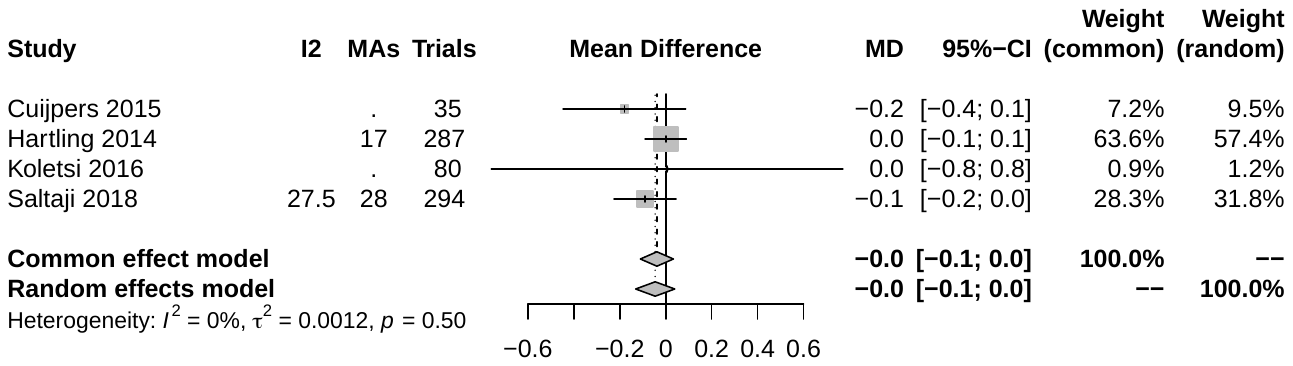

#### Supplemental figure 4.9. Forest plot of patient and healthcare provider blinding for continuous outcomes in double-blind vs open-label studies.

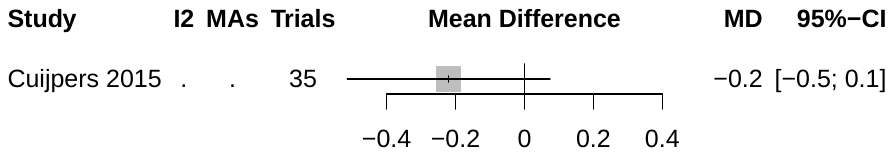

#### Supplemental figure 4.10. Forest plot of patient and healthcare provider blinding (adjusted) for continuous outcomes in double-blind vs open-label studies.

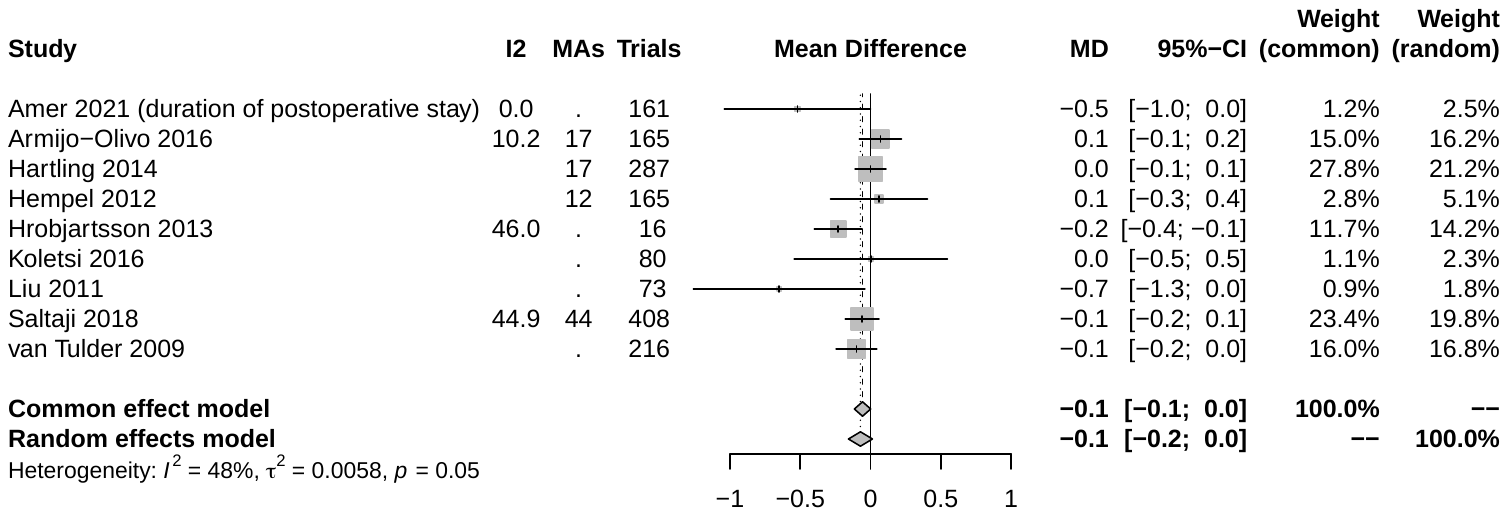

#### Supplemental figure 4.11. Forest plot of outcome assessor/adjudicator blinding for continuous outcomes in double-blind vs open-label studies.

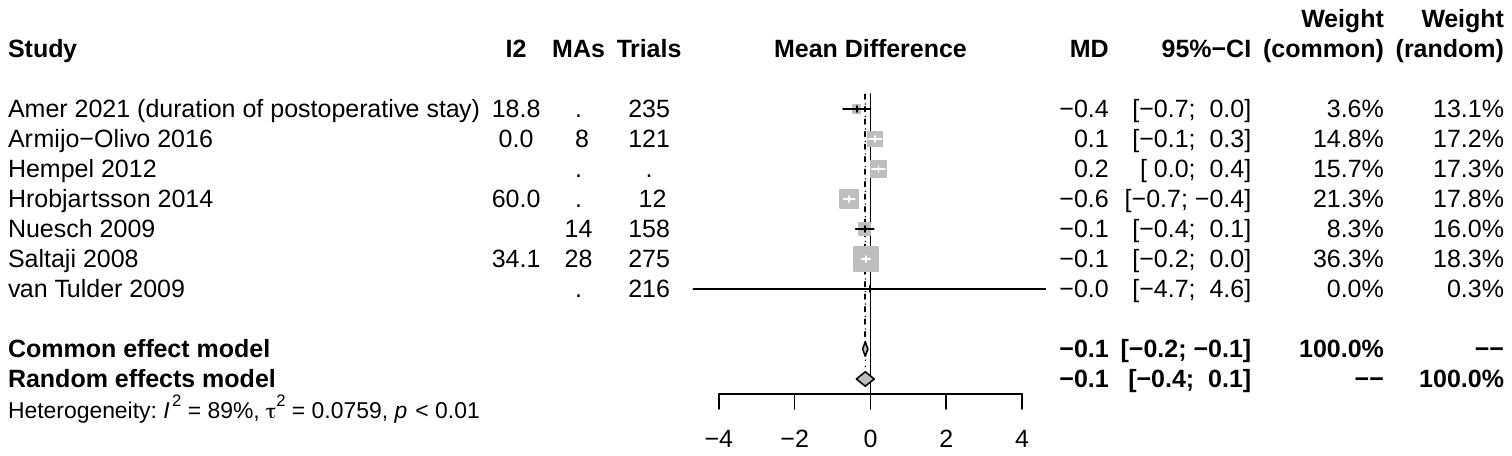

#### Supplemental figure 4.12. Forest plot of patient blinding for continuous outcomes in double-blind vs open-label studies.

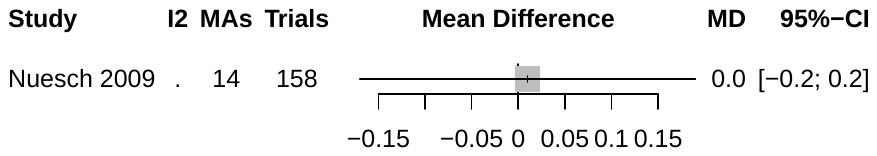

#### Supplemental figure 4.13. Forest plot of patient blinding (adjusted) for continuous outcomes in double-blind vs open-label studies.

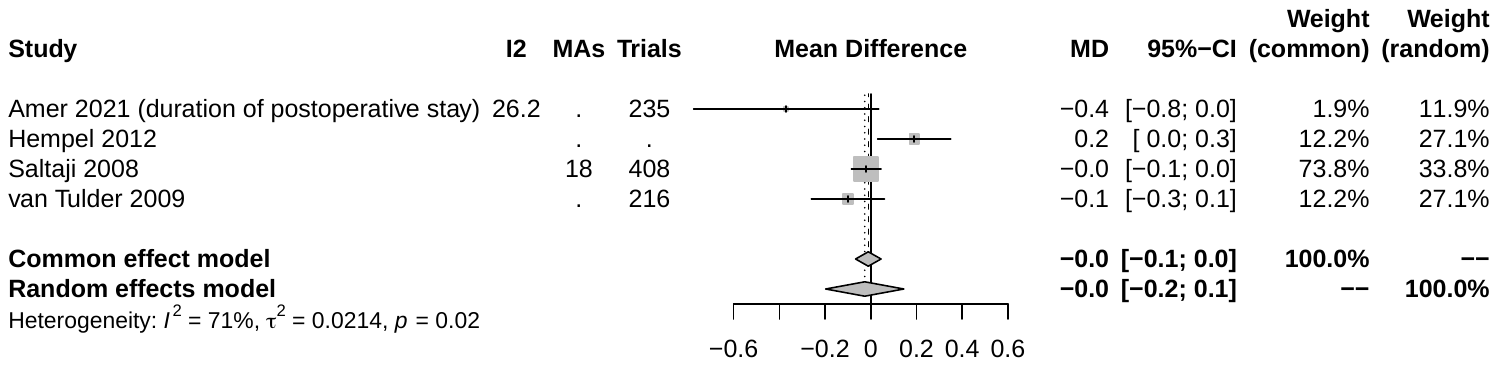

#### Supplemental figure 4.14. Forest plot of healthcare provider blinding for continuous outcomes in double-blind vs open-label studies.

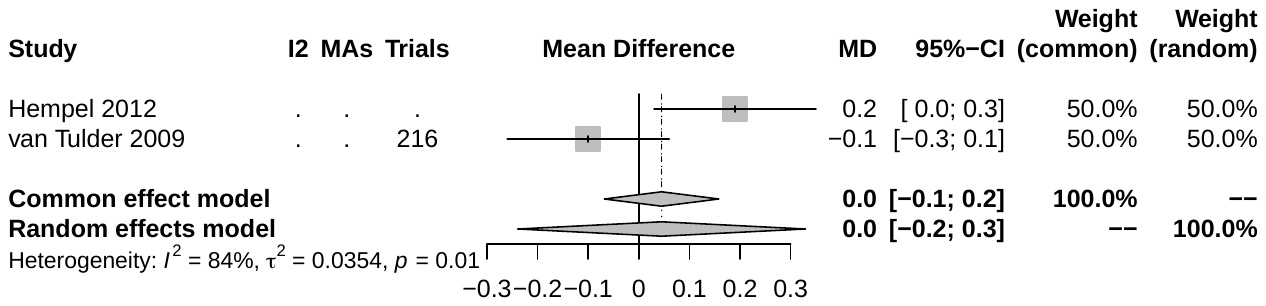

#### Supplemental figure 4.15. Forest plot of healthcare provider blinding (adjusted) for continuous outcomes in double-blind vs open-label studies.
